# The optics of the human eye at 8.6 µm lateral resolution

**DOI:** 10.1101/2021.02.26.20241604

**Authors:** Sergio Bonaque-González, Juan M. Trujillo-Sevilla, Miriam Velasco-Ocaña, Óscar Casanova-González, Miguel Sicilia-Cabrera, Alex Roqué-Velasco, Sabato Ceruso, Ricardo Oliva-García, Javier Martín-Hernández, Oscar Gomez-Cardenes, José G. Marichal-Hernández, Damien Gatinel, Jack T. Holladay, José M. Rodríguez-Ramos

## Abstract

Ocular optics is normally estimated based on 2,600 measurement points within the pupil of the eye, which implies a lateral resolution of approximately 175 microns for a 9 mm pupil diameter. This is because information below this resolution is not thought to be relevant or even possible to obtain with current measurement systems. In this work, we characterize the in vivo ocular optics of the human eye with a lateral resolution of 8.6 microns, which implies roughly 1 million measurement points for a pupil diameter of 9 mm. The results suggest that the normal human eye presents a series of hitherto unknown optical patterns with amplitudes between 200 and 300 nm and is made up of a series of in-phase peaks and valleys. If the results are analysed at only high lateral frequencies, the human eye is also found to contain a whole range of new information. This discovery could have a great impact on the way we understand some fundamental mechanisms of human vision and could be of outstanding utility in certain fields of ophthalmology.

## Introduction

Knowledge of the optics of the human eye has allowed us to broaden the understanding of the mechanism of human vision. It has led the current field of ophthalmology to a highly technical level where customized treatments and instrumentation-based diagnostics have become the norm. However, measuring the optics of the eye is not a trivial task. In 1961, Smirnov developed an advanced version of the Scheiner disk [1] to subjectively estimate the optical imperfections of the eye, also called aberrations or phase map, in a process named ocular aberrometry [2]. At that time, he anticipated that his invention would have no practical application, stating that “…the calculations take 10–12 hours […] Therefore, it is unlikely that such detailed measurements will ever be adopted by practitioner-ophthalmologists.” He did not foresee that advances in computer science would greatly speed up calculations and that scientists would find the measurement of ocular aberrations an invaluable tool, progressing from a research instrument to a clinical application system. Indeed, some of modern ophthalmology could not be understood without the deep understanding of ocular optics that aberrometers have made possible.

Currently, there are several options for the estimation of ocular aberrations based on different techniques, i.e., Hartmann-Shack sensors (H-S) [3], pyramidal sensors (P-S) [4], interferometric techniques [5], laser ray tracing [6] and curvature sensors [7]. Techniques based on H-S sensors are the most widely used in ophthalmology. However, they are restricted to sampling the phase map at up to 2,600 measurement points within the pupil of the eye (approximately 175 µm of lateral resolution for a 9 mm pupil diameter [8]) and also suffer from a limited dynamic range [9]. These limitations constrain its usefulness in abnormal eyes, which are precisely the most interesting to characterize [10]. Currently, P-S offers the highest resolution in ocular aberrometry, up to 45,000 measurement points (approximately 37 µm theoretical lateral resolution for a 9 mm pupil diameter) [8]. Nevertheless, P-S generally suffers from non-linear behaviour, diffraction effects between its different pupil images, and a relatively limited dynamic range as a result of the trade-off between the slope accuracy and achievable spatial resolution [11]. These drawbacks cause the phase maps obtained to be somewhat fuzzy, and the details that can be visualized do not match the theoretical resolution [8]. Therefore, the H-S sensor is still considered the gold standard in ophthalmology.

Actual knowledge about the optics of the normal human eye can be roughly summarized as follows: it has relatively smooth optics with no optical structures that diverge too greatly from a conical shape with some astigmatism. Its phase map is usually expressed as a decomposition of orthonormal polynomials within a circular pupil, in particular Zernike polynomials [12], where most of the deformations can be described with the first two to three dozen terms [13, 14]. Thus, up to 66 Zernike terms are usually considered sufficient to represent ocular optics with high quality even in pathological eyes, assuming a loss of high frequencies that is not considered to hide any relevant information. [15, 16]. However, some authors have already drawn attention to the inefficiency of this approach [17].

In this work, we characterize the ocular optics of living human eyes with a lateral resolution of approximately 8.55 µm. This implies more than a million points of measurement for a pupil with a diameter of 9 mm, orders of magnitude higher than the current gold standard offered by the H-S sensor. We show evidence that when ocular optics are measured at a sufficiently high resolution, a series of phase structures emerge. We hypothesize that this finding could have a great impact on some current ophthalmic surgical procedures and on the clarification of some fundamental mechanisms of human vision.

## Methods

### The phase sensor

To measure the phase map, we use the wavefront phase imaging (WFPI) sensor, which offers high resolution and has been used with success in other fields, such as semiconductor silicon metrology [18]. WFPI captures two intensity images around the pupil plane of the instrument using a standard image sensor, i.e., a charge-coupled device (CCD). Then, the phase gradients along two orthogonal directions are recovered from both the captured intensity distributions. A standard WFPI setup for transparent samples is depicted in figure 1a. A collimated beam (with an almost flat phase map) from a light-emitting diode (LED) is used as the light source. The use of incoherent light, in contrast to the H-S sensor, makes this sensor insensitive to speckle effects. The light beam passes through the sample, which modifies the shape of the beam according to the optical behaviour of the sample, thus generating a distorted outgoing phase map. An imaging telescope composed of a pair of converging lenses, *L*_1_ and *L*_2_, translates the plane of the sample, *P*, into a conjugated plane, *P*’. The two required intensity images, *I*_1_ and *I*_2_, are acquired around *P*’ in equidistant planes along the z-axis. Choosing focal lengths of *L*_1_ and *L*_2_ appropriately allows us to adjust the sample size to the sensor size. This optical setup is similar to some proposals for curvature sensors [7]; however, the mathematical treatment of the data in WFPI is completely different, as explained below.

**Figure 1.**
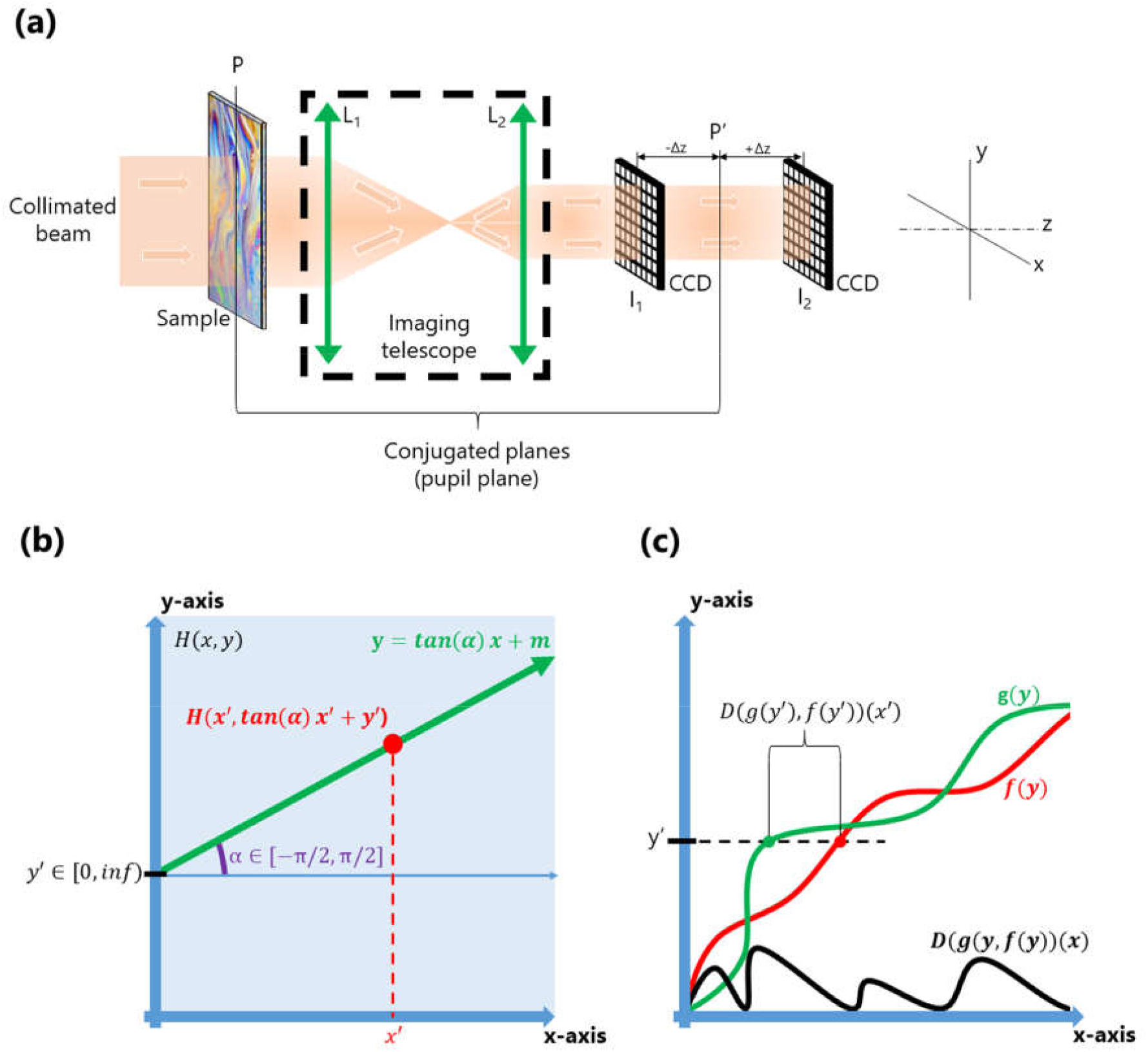
(a) Basic optical setup of the wavefront phase intensity (WFPI) technique for transparent samples; (b) graphical representation of Eq. 1; (c) graphical representation of Eq. 2. P: pupil plane; L: lens; I: intensity image; CCD: charge-coupled device; P’: conjugated pupil plane.

Let *H*(*x, y*) be a continuous two-dimensional function defined for positive values of *x, y* and taking only positive numbers as values, and let *V* be an auxiliary transformation function acting on *H* as follows:

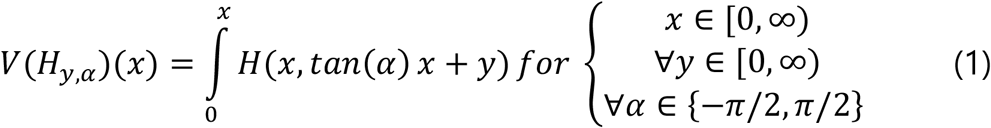

where *α* is a variable composed of a set of *k* discrete angles uniformly distributed in the interval [−*π*/ 2, *π*/ 2], each defining a line with its origin in *y* and an angular separation from the *x* − *axis* equal to a (see figure 1b). Then, for each pair of values *y* and α, *V*(*H*_*y, α*_)(x) is a one-dimensional function that represents the accumulated *H*(*x, y*) values along the defined line of *H*. The graphical representation of *V*(*x*) for particular values of (*y, α*) is similar to one of the coloured curves, green or red, in figure 1c.

Let *D* be another auxiliary function that represents the distance on the abscissa axis between two unidimensional continuous functions represented by *f*(*x*) and *g*(*x*), defined for positive values of *x* and yielding only positive numbers. Figure 1c shows a graphical representation of *D*, which can be mathematically described as:

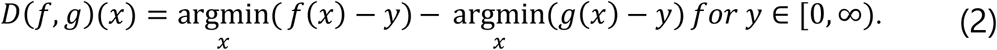

Let *I*_1_(*x, y*) and *I*_2_(*x, y*) be two continuous bi-dimensional functions representing two different intensity maps from which it is desired to recover the phase map. The phase gradients in the horizontal and vertical directions, *φ*_*h*_ and *φ*_*v*_, are directly related to *I*_1_and *I*_2_ as follows:

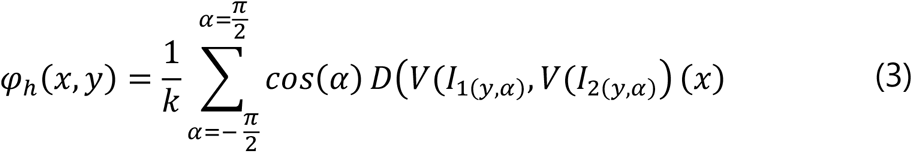

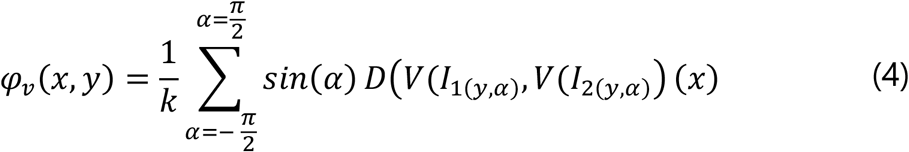

where *k* is the number of discrete angles considered, defined as:

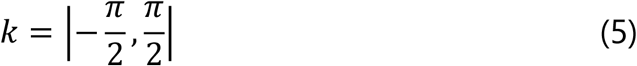

Note that the units of *φ*_*h*_ and *φ*_*v*_ correspond to those of the input intensity images, i.e., pixels. To convert these units into physical units, i.e., metres, we apply a factor that takes into account the pixel size, *s*, and the distance between the images, 2Δ*z*:

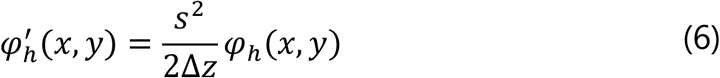

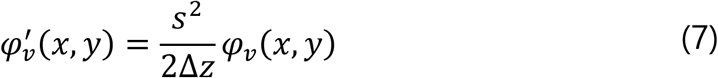

Finally, numerical integration is required to obtain the phase map from its derivatives. In our case, we use the algorithm developed by Talmi & Ribal [19]; however, very similar results can be obtained using other methods [20].

For simplicity, the above mathematical equations have been made assuming continuous functions, but the same reasoning can be followed for a real implementation where *I*_1_and *I*_2_ are discrete and finite intensity image maps. In this case, it would be necessary to apply affine transformations to functions *D* and *V* to be able to deal with the discrete nature of the real intensity images, which can be done computationally in an intuitive way through interpolation. Regarding the value of *k*, we did not find a significant increase in the precision of the algorithm when *k* > 120 for resolution ≤ 4096 × 4096 and pixel size ≈ 9µ*m*. However, with other configurations, the optimal value of *k* should be studied. This is a generalized and formal explanation of the WFPI basis, and its computational implementation is simpler. We encourage the interested reader to consult the step-by-step explanation of the algorithm in patent PCT/EP2018/052533 [21], which is more focused on the computational and discrete aspects of the methodology.

### The apparatus

Measurements were performed in a custom-developed double-pass optical setup incorporating a WFPI implementation. Its current configuration, outlined in figure 2, has 4 different channels:

**Figure 2.**
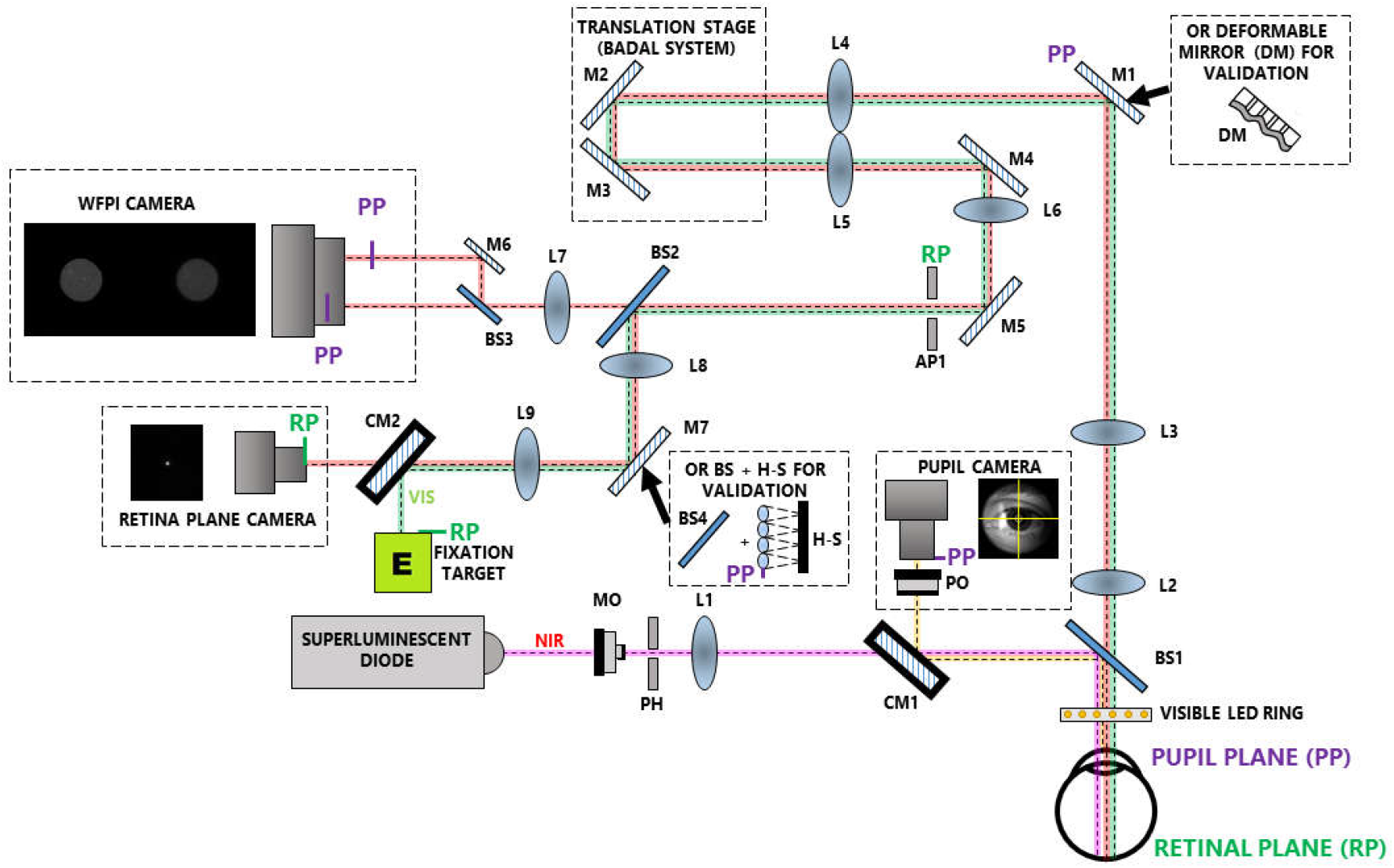
Schematic diagram of a custom-made optical system with 4 different channels: (1) the illumination channel (pink line); (2) the pupil monitoring channel (yellow line); (3) the science channel (red line); and (4) the fixation target channel (green line). NIR: near infrared light; MO: microscope objective; PH: pinhole; L: lens; CM: cold mirror; PO: photographic objective; BS: beam splitter; M: mirror; AP: aperture; H-S: Hartmann-Shack sensor; VIS: visible light.

1. The illumination channel contains a single-mode superluminescent diode (SLD) (QSDM-780-15D, QPhotonics, LLC, Ann Arbor, USA) with a central wavelength (*λ*) of 780 nm and a spectral bandwidth of 20 nm. It works in combination with a control unit (CLD1015, Thorlabs, Newton, USA) that allows the maximum emission time to be set at 35 ms. The microscope objective focuses light into a pinhole that removes aberrant rays. Then, an achromatic lens is placed with its back focal point in the pinhole to obtain a collimated (nearly parallel) beam. After passing through a cold mirror and a non-polarizing 10:90 beam splitter, the beam is directed towards the eye with a beam diameter of approximately 800 µm. An offset of the beam from the centre of the pupil is introduced to avoid corneal reflections in the WFPI intensity images. The maximum permissible exposure in the pupil plane for 35 ms at *λ*=780 nm is 2.4 mW, which is calculated using the safety limits of the ANSI standards for the wavelength used [22]. We measured 0.78 mW in the pupil plane, which is well below the ANSI safety standards.
2. The pupil monitoring channel allows the position of the subject to be monitored during the measurements. It consists of a monochrome camera (UI-3370SE-M-GL, IDS, Obersulm, Germany) in combination with a camera lens that conjugates the pupil plane of the system with that of the camera. A ring with 18 small LEDs of *λ*=630 nm placed before the eye provides the illumination for this channel. Its light power measured in the corneal plane is orders of magnitude below the ANSI standard safety limits for the wavelength used.
3. The science channel. The retina spreads the incoming light from channel 1 from back to front. When the dispersed light beam emerges from the eye, it will carry the phase information for the entire eye, including the cornea and the lens. This beam is guided through a variety of converging lenses to adjust the beam size to the WFPI camera size and to correct for defocus in the Badal system in a range of [-10.00 +10.00] D. As the presence of defocus will change the size of the intensity images in the WFPI, its correction by the Badal system allows us to provide an almost constant lateral resolution between eyes with different spherical refractions. A monochromatic camera positioned in the retinal plane (UI-3370SE- M-GL, IDS, Obersulm, Germany) allows us to estimate the Badal position that offers the best spherical correction by maximizing the central intensity of the focused light beam. In the event of an error in the correction process of the Badal system, it is shown by specific measurements that residual defocus values can be characterized by the WFPI sensor in a range of ±6.00 D. An aperture is placed in a retina plane to reduce reflections within the system. To capture two intensity images around the pupil plane of the eye at a separation of 13.33 mm, a small system composed of a 50:50 beam splitter and a mirror divides the science beam into two parts, which are simultaneously captured by a low-noise, 11-megapixel cooled camera (Bigeye G-1100 Cool, Allied Vision Technologies, Stadtroda, Germany). The magnification of the apparatus is approximately 1.0523, and each pixel in the estimated phase map represents approximately 8.553 µm in the plane of the pupil of the eye.
4. The fixation target channel consists of a modified liquid crystal display (LCD) retro-illuminated monochromatically at *λ*=620 nm. The projected image is a binary Maltese cross that can be more clearly observed by the experimental subject once the Badal system has made the spherical correction. The centre of the cross is coincident with the optical axis of the system, so that when the subject fixes on it, the optical axis of the system is aligned with the line of sight. Its light power measured in the corneal plane is orders of magnitude below the ANSI standard safety limits for the wavelength used.

For calibration and validation of the system (see next section), an H-S sensor (WFS20-5C, Thorlabs, Newton, USA) and */* or a deformable mirror (DM) (DM140A-35-UP01; Boston Micromachines, Boston, USA) were placed in the pupil planes within the system, as depicted in figure 2.

The optical setup was mounted on an x-y-z translation stage that was manually controlled by a joystick to align the pupil of the eye with the instrument using the pupil-monitoring channel. A conventional ophthalmic chinrest intended to position the experimental subject was attached to the table that supported the entire instrument.

All the optoelectronic and mechanical elements of the apparatus were automatically controlled and synchronized using custom-built software in C++. All processing of the H-S and WFPI intensity images was performed using custom-built software in MATLAB (v2019a, MathWorks Inc., Natick, USA).

The WFPI software provides the phase map within the actual pupil of the measured element, which can be circular or not. When measuring at a high resolution, the fit to the Zernike polynomials does not provide a good physical representation of the object under study [17].

Nevertheless, in all cases, a Zernike adjustment was made for compatibility with the H-S sensor used for validation and to split the overall phase error into the main Zernike components. The required circular pupil defined for this fitting was the largest one inscribed within the real pupil when not specified otherwise.

### Calibration and validation

For the calibration of the prototype, a custom-designed eye model, composed of an artificial retina rotating at 300 rpm, an artificial cornea (an achromatic doublet with a focal length of 40 mm), and an artificial pupil with a 7 mm diameter, was placed in the measurement position. All the components of the eye model shared the same optical axis. Its emmetropic condition, when needed, was calibrated with a shearing interferometer (SI100, Thorlabs, Newton, USA). A series of defocuses was induced by increasing or decreasing the distance between the retina and the cornea of the artificial eye, in a range of approximately −7.00 to 7.00 D in 13 steps. The defocus status of the artificial eye was determined by independent measurement (outside the apparatus) with the H-S sensor using a small optical assembly designed for this purpose. Additionally, cylindrical trial lenses of +0.50, +1.00, +2.00 and +3.00 D with axes of 90, 45, and 22° were centred in front of the emmetropic artificial eye with back vertex distances of 5 mm. The measurements were recorded with respect to the pupil plane and compared to the powers of the trial lenses, transferring the obtained power from the pupil plane to the trial lens plane. For both conditions, defocus and astigmatism, the phase map was estimated using the WFPI sensor by taking 3 consecutive measurements. The defocus or astigmatism in diopters was calculated from the obtained phase maps by decomposing it into Zernike polynomials and using the methodology proposed by Salmon T. et al. [23], with the difference that half of the cylindrical value was not added to the value of the sphere to facilitate the determination of the origin of possible errors.

The repeatability of the instrument was tested by using the “continuous”-mode capturing option of the science camera, which acquires an intensity image approximately once per second (even though the exposure time can be much shorter). One hundred consecutive images were taken with the eye model with a 7 mm pupil diameter in which a slight fixed defocus was induced by manually moving the position of the retina. For the reproducibility tests, we made measurements under the same conditions, but two technicians took turns performing the measurements alternately, misaligning and realigning the device between each measurement. The results are reported in terms of the root-mean-square (RMS) error. Additionally, each individual phase map was decomposed into the first 66 Zernike polynomials according to the OSA standards [24] without considering the piston or the tip / tilt. The analysis in terms of the RMS error is shown for low-order (Zernikes 3 to 5), medium-order (Zernikes 6 to 14), and high-order (Zernikes from 15 to 65) aberrations.

For validation in the presence of higher frequencies (high-order aberrations), a DM was placed in a pupil plane (see figure 2) and directed with 800 phase maps with amplitudes uniformly ranging between 0.35 and 2.8 µm in 0.35 µm steps. The phase maps that fed the DM were composed of the random values of the first 66 Zernike polynomials (piston and tip / tilt were not included). For this test, the H-S sensor was placed in a pupil plane within the setup (see figure 2) to simultaneously estimate the phase. The pupil diameter was restricted to 4 mm due to the small size of the aperture of the DM (4.4 mm). To make comparisons between the sensors possible, the phase maps obtained by both sensors were resampled to the DM input resolution by bilinear interpolation. The images in each measurement were bias-subtracted and flat-fielded (with the DM in its resting position). The results in terms of RMS errors were statistically analysed in Microsoft Excel 2016 (Microsoft, Redmond, WA) by means of the two-tailed Student’s t-test (two-sided).

### In vivo tests

Finally, five living human eyes belonging to five subjects aged between 27 and 38 years were tested monocularly with the apparatus and with the H-S sensor placed in the system, as depicted in figure 2, in order to simultaneously obtain a reference map. The exposure time for each of the measurements was 30 ms. First, the repeatability/reproducibility in a real eye was estimated by taking 20 “consecutive” measurements, allowing the volunteer to blink between measurements, with a time between measurements of approximately 3 seconds. Second, simple measurements were taken with the apparatus for the remaining 4 eyes. All the eyes tested had a best corrected visual acuity of 20/20 or better and had no observable pathology. These eyes were not chosen for having any specific characteristics and correspond to volunteers from the research laboratory itself. As the instrument is capable of taking images in 30 ms, cycloplegia was not used, and nor was a bit-bar required to take the measurements. The images in each measurement were bias-subtracted and flat-fielded. The subjects were aligned with the instrument using the x-y-z stage, while the natural pupil was viewed on a monitor using the pupil camera. As in the previous case, a simple estimation of the sphero-cylindrical correction was carried out using the same methodology as described above.

When the results of both sensors, the WFPI and H-S, were compared, the phase map obtained by WFPI was resampled to the H-S resolution by bilinear interpolation, and both maps, the H-S and WFPI, were decomposed into the first 66 Zernike polynomials. Since the pupils of real eyes are not perfectly circular, the H-S edge microlenses and their corresponding areas in WFPI were removed from processing to ensure the measurement of an area completely covered by the microlenses.

This study was approved by the Ethics Committee of the Hospital Universitario de Canarias, was conducted in accordance with the provisions of the Declaration of Helsinki, all volunteers were acquainted with the nature and possible consequences of the study, and provided written informed consent. Also, all subjects gave their consent for the publication of the data.

## Results

### Calibration and validation

The apparatus was able to recover the defocus introduced by the artificial eye with good precision, as shown in figure 3a. The error bars are not visible because the standard deviation was very low in all cases. The mean error was 0.04 ± 0.06 D (range [0.003 0.12]). Regarding the cylinder, introduced by means of +0.50, +1.00, +2.00 and +3.00 D cylindrical trial lenses with 90°, 45°, and 22° axes, the results are shown in figure 3b for all the measurement conditions. The mean absolute error in the cylinder power was 0.05 ± 0.05 D (range [0 0.18]). The worst measurements were clearly those corresponding to the 2.00 D cylinder oriented at 90 degrees (error of 0.18 ± 0.005 D). We do not know why these specific measurements were not as accurate as the rest of the measurements at 90 degrees or with the 2.00 D lens. The mean error in the axis orientation was 1.67 ± 2.06 degrees (range [0 6.6]). This seems reasonably accurate, as the trial lenses were oriented following a manual procedure using the pupil camera as a reference. Small angular differences between the science camera and the pupil camera could also explain this apparent inaccuracy.

**Figure 3.**
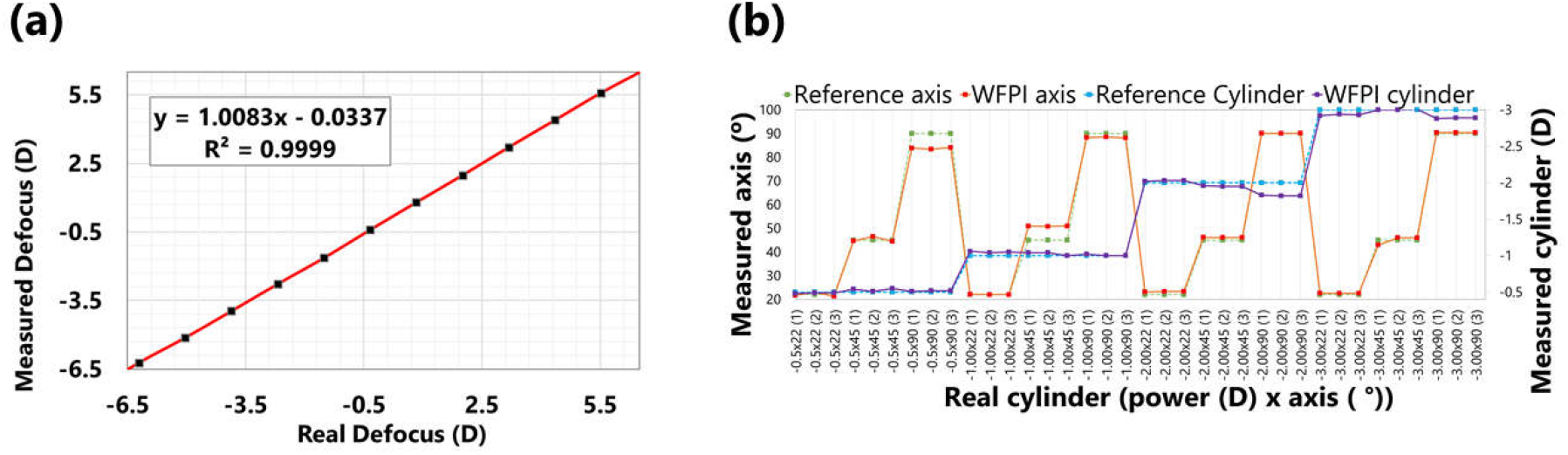
Calibration of the apparatus for different induced defocuses and astigmatisms. (a) Defocus induced in artificial eyes with different axial lengths versus defocus recovered by the apparatus. (b) Different cylindrical trial lenses in different orientations and their measurement with the apparatus. Three different measurements were taken in each condition, and the data were for a 7 mm pupil diameter.

Calculations of the repeatability were performed using 100 consecutive measurements on an artificial eye with a 7 mm pupil diameter and yielded an average phase RMS of 0.984±0.014 µm, with a maximum RMS difference between any two phase maps of 0.063 µm. When the obtained phase maps were decomposed into the first 66 Zernike polynomials, the low-order RMS (Zernikes 3 to 5) was 0.57±0.008 µm, with a maximum difference of 0.037 µm. The medium-order RMS (Zernikes 6 to 14) was 0.051±0.004 µm, with a maximum difference of 0.019 µm. The high-order RMS (Zernikes 15 to 65) was 0.01±0.001 µm, with a maximum difference of 0.005 µm.

The reproducibility calculations performed on 100 measurements on an artificial eye with a 7 mm pupil diameter yielded an RMS of 1.024±0.028 µm, with 0.129 µm being the maximum difference between any two phase maps. When the obtained phase maps were decomposed into the first 66 Zernike polynomials, the low-order RMS (Zernikes 3 to 5) was 0.599±0.02 µm, with a maximum difference of 0.092 µm. The medium-order RMS (Zernikes 6 to 14) was 0.055±0.008 µm, with a maximum difference of 0.036 µm. The high-order RMS (Zernikes 15 to 65) was 0.011±0.002 µm, with a maximum difference of 0.009 µm.

Figure 4 shows the validation of the apparatus in the presence of high frequencies induced by a DM for a 4 mm pupil diameter. The RMS error was calculated for each of the measurements by subtracting the phase obtained with each sensor from the reference map. To make this operation possible, each measured phase map was resampled to the resolution of the reference map using bilinear interpolation. As shown in figure 4a, both sensors, the H-S and WFPI, showed similar behaviour, with the WFPI being statistically (p<0.05) more precise for amplitudes between 0.35 and 1.75 µm. No significant outliers were observed in either of the two sensors. Small errors with respect to the reference map may be due to the resampling process by interpolation, or they may have a component associated with inaccuracies in the characterization of the response of the microactuators for complex input maps [25]. In figure 4b, the first row shows the case in which the WFPI sensor yielded its best result (amplitude or peak to valley (P-V) 2.45 µm; RMS errors of 0.036 and 0.076 µm for the WFPI and H-S sensors, respectively), while the second row shows its worst result (P-V 2.1 µm; RMS errors of 0.202 and 0.124 µm for the WFPI and H-S sensors, respectively). The third row shows the case in which the H-S sensor gave its worst result (P-V 1.75 µm; RMS errors of 0.048 and 0.167 µm for the WFPI and H-S sensors, respectively). We do not show the best case for the H-S sensor, since WFPI also showed a very good result (P-V 1.05 µm; RMS errors of 0.047 and 0.032 µm for the WFPI and H-S sensors, respectively). The first column of figure 4b shows the result of the WFPI sensor at full resolution. When measured at a high resolution, a mesh-like lattice emerges as a background image, as seen in figure 4b. This background is compatible with the shape of the microactuators, so their individual influences on the phase can be estimated.

**Figure 4.**
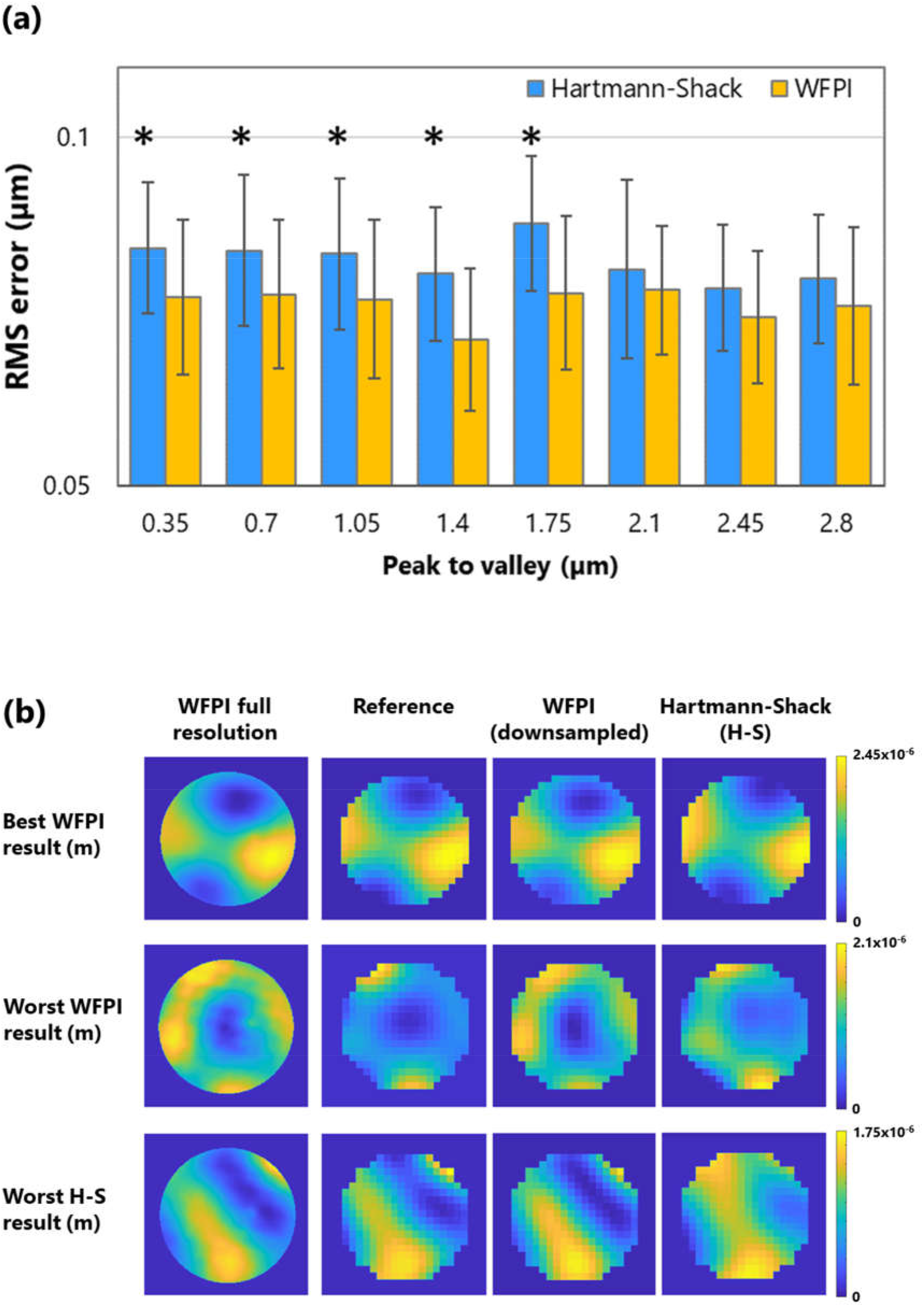
Instrument validation results using the WFPI sensor, a deformable mirror (DM), and a Hartmann-Shack sensor (H-S). (a) For a variety of different amplitudes or peak-to-valley values, the average root-mean-square (RMS) errors of the residual phase map calculated by subtracting the measured phase maps from the reference maps and the error bars corresponding to the standard deviation are shown. An asterisk, *, indicates a statistically significant difference between the two sensors (p-value <0.05). (b) The first row shows the case in which the WFPI sensor yielded its best result, the second row shows the case in which the WFPI sensor yielded its worst result, and the third row shows the case in which the H-S sensor yielded its worst result. The first column shows the high-resolution result of the WFPI sensor. The data are for a 4 mm pupil diameter.

### In vivo test

As a first step, repeatability/reproducibility was estimated in a real eye. It was a myopic eye with a spherical equivalent of approximately −6.21 D (eye #MVO in figures 5 and 6) and a naturally unusually large pupil size. 20 relatively consecutive measurements were taken. The subject was instructed to blink between measurements, with the approximate time between each being 3 seconds. As the measurements were performed without pharmacological dilation of the pupil, its diameter ranged from 7.3 to 7.7 mm during the test.

**Figure 5.**
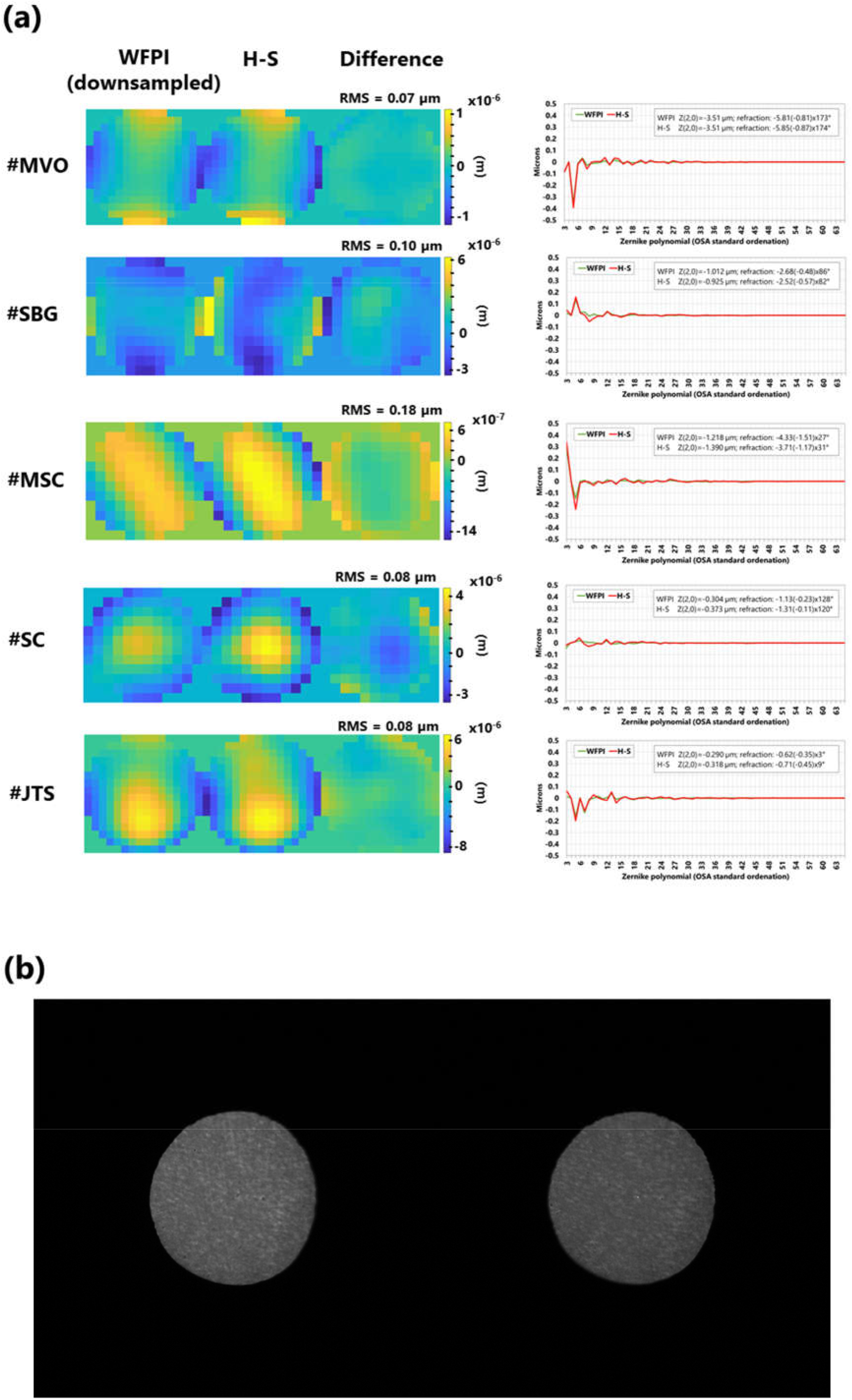
Measurements of 5 real eyes with the device. (a) Recovered phase map with the WFPI and with the H-S as well as its difference map together with its RMS value. On the right, the comparison of Zernike polynomials can be seen. For clarity, the defocus value (Zernike 4) is set to zero on the graph, and the numerical value is provided, along with the sphero-cylindrical refraction, in the upper box of each individual graph. (b) Example of intensity images captured with the WFPI sensor, corresponding to eye #MVO.

**Figure 6.**
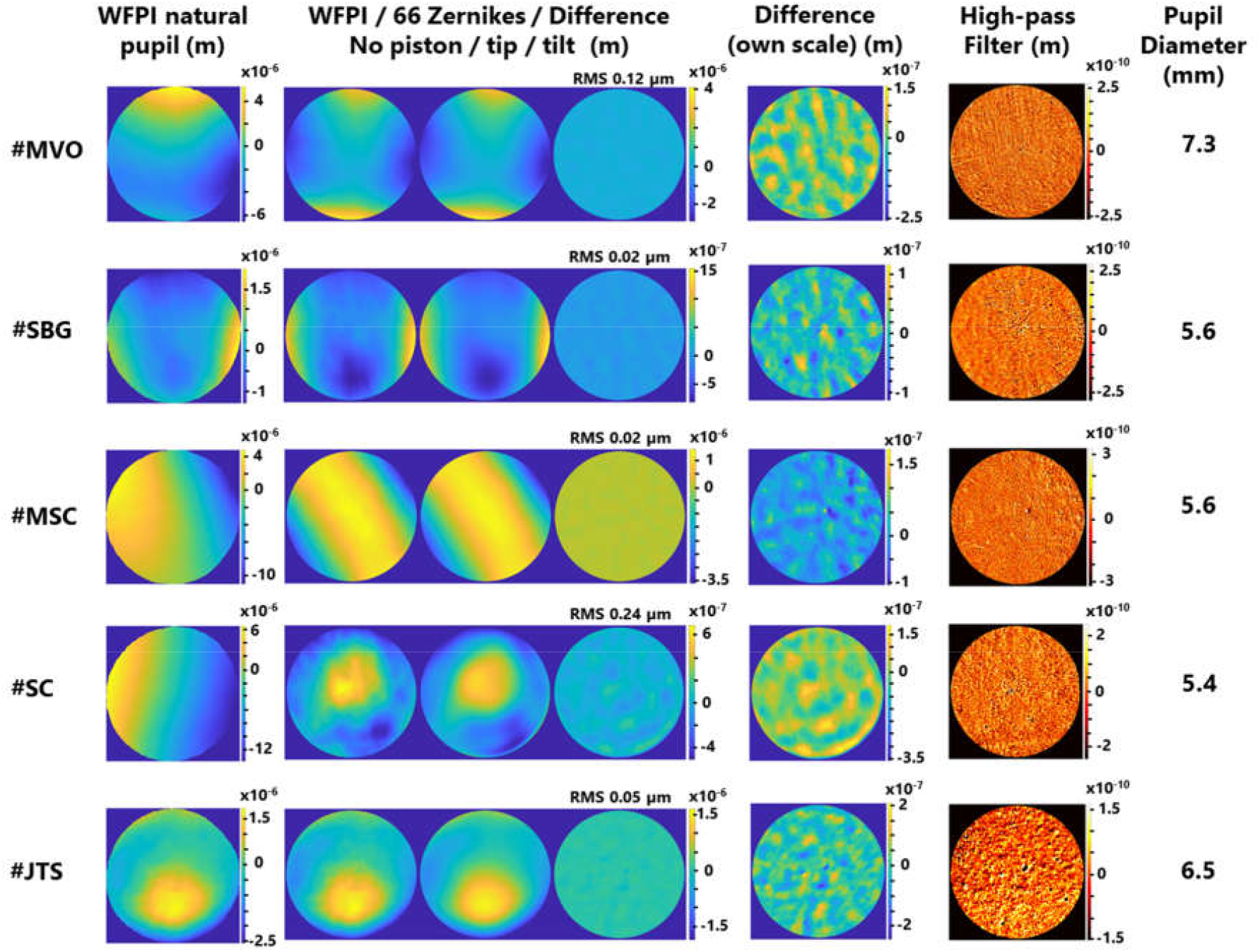
Measurements of 5 real eyes (the same as in figure 5) with the device and its analysis with a lateral resolution of 8.55 microns. The first column is the phase map retrieved by the apparatus. The second column is a group of 3 sub-images comprising the WFPI result within a circular pupil and without piston/tip/tilt, its fit to the first 66 Zernike polynomials, and the difference between the two. The third column is the same difference phase map as above but with its own colour map or scale. The fourth column is the first phase map of the second column, to which a Gaussian filter with sigma = 0.5 was applied, thus revealing the high-frequency components of the phase. Finally, the last column indicates the diameter of the pupil when considering a circular pupil (the largest inscribed pupil).

Therefore, the analysis was restricted to a pupil diameter of 7.3 mm for all measurements. Piston and tip/tilt were removed from the measurements. The analysis of all obtained phase maps yielded a total RMS of 11.028±0.32 µm, with the maximum difference between any two phase maps being 1.25 µm. When the obtained phase maps were decomposed into the first 66 Zernike polynomials, the low-order RMS (Zernikes 3 to 5) was 6.198±0.196 µm, with a maximum difference of 0.801 µm. The medium-order RMS (Zernikes 6 to 14) was 0.109±0.01 µm, with a maximum difference of 0.034 µm. The high-order RMS (Zernikes 15 to 65) was 0.012±0.002 µm, with a maximum difference of 0.013 µm. These measurements can be understood as halfway between repeatability and reproducibility since accommodation is not paralyzed. Additionally, its activation could explain the variability in the total RMS and in the pupil diameter.

Figure 5a shows the comparative measurements of both sensors in the apparatus, the WFPI and H-S, for the 5 different real eyes, both as phase maps together with a difference map and in terms of the Zernike polynomial decomposition. In the latter, the defocus value (Zernike 4 or Z(2,0)) was set to zero to allow a better visualization of the graph, as defocus was the predominant component in almost all the eyes. Nevertheless, its value is shown in the data included in the graph, which also shows the sphero-cylindrical refraction of the eyes calculated from the data of both sensors. To make these comparisons, the WFPI measurement in circular pupils was resampled at the resolution of the H-S sensor. The differences in RMS between the two sensors were on the order of those observed during the device validation process. However, in eye #MSC, a clinically significant difference (>0.25 D) in refraction was detected in both the cylinder and defocus. We do not have a clear explanation of why this difference occurred, although we verified that our custom software for H-S data treatment detects high oblique astigmatisms poorly. Figure 5b shows an example of what the intensity images captured by the instrument look like, corresponding to eye #MVO with a pupil diameter of 7.3 mm. Although we have not verified this, we believe that this type of intensity image could be useful in the diagnosis or monitoring of ocular pathologies that modify the amplitude, such as the presence of cataracts.

All of the above leads us to figure 6, which is the main result of this work. These are the same eyes and measurements as those displayed in figure 5, but the entire phase maps were analysed in depth, making use of the whole available lateral resolution, approximately 8.55 µm. The first column, labelled “WFPI natural pupil”, shows the measurement result as it is returned by the apparatus. Note that this measurement was performed on the subject’s natural pupil, which in no case turned out to be completely circular. As these measurements include the piston and tip/tilt, the details are masked by their high amplitudes. The second column, labelled “WFPI / 66 Zernikes / Difference”, is made up of 3 sub-figures. The first sub-figure shows the previous result but within a circular pupil (the largest circular pupil inscribed within the natural pupil). In addition, the piston and tip/tilt have been removed. The second subfigure shows the fit to the first 66 Zernike polynomials. The third sub-figure shows the difference between the two previous phase maps together with its RMS. The third column, labelled “Difference (own scale)”, shows the difference between the phase map adjusted to Zernikes and without the adjustment with its own colour map, that is, the third sub-figure of column 2 but with its own scale. This last image represents the information that cannot be captured with a decomposition in terms of the first 66 Zernike polynomials (similar to the information that is usually provided by commercial aberrometers) and is the main contribution of this work. In all cases, a substructure of peaks and valleys emerges with an approximate amplitude between 200 and 300 nm. The case of eye #SC is remarkable, as there were 0.24 microns of RMS that could not be characterized with a decomposition in terms of the first 66 Zernike polynomials, which is a significant quantity for the human eye. Finally, the last column, labelled “High-pass filter”, shows the result of the first sub-image of the second column, to which a Gaussian filter of sigma 0.5 was applied to reveal the high-frequency components of the phase map. There were notable differences between the 5 eyes. While some sort of vertical line is observed in eye #MVO (the most myopic), this pattern is not present in the rest of the eyes. Although we cannot be sure of the origin of these lines, they are compatible with the molecular arrangement of some macular components, such as the xanthophilic pigment of the macula lutea, which is arranged in parallel lines [26], and this could have somehow been transmitted to the recovered phase map. Its origin could also be originated in tensions of the posterior capsule. Future studies will be necessary to clarify the origin of these lines, since we have already ruled out instrumental effects. In eye #SBG, a filamentous structure with an amplitude on the order of 0.4 nm can be seen that would be compatible with a floater in the vitreous body. Eye #JTS, the most emmetropic, is perhaps the most striking due to its differences with respect to the rest, since a mottling can be observed that covers the entire pupil. We hypothesize that this finding may be related to some degree of dry eye. Applying high-pass filters is somewhat tricky, and different filters can be applied that reveal different phase components. We applied this filter specifically because we think it reveals the underlying structure well in most eyes. However, for the interested reader, we provide the raw phase maps as additional data (column 1 of figure 6). We encourage alternative analyses.

## Discussion

In this work, we provided strong evidence that ocular optics are more complex than previously assumed and, below the previous resolution limits, there is a large amount of information that until now has been unknown. To visualize the optics with a lateral resolution of approximately 8.55 µm, we used a relatively new phase sensor, the WFPI sensor. An exhaustive calibration and validation of the WFPI with the apparatus was carried out, focusing on ocular measurements, using all the tools at our disposal, and improving (by including more tests) the methodology used for the validation of other ocular aberrometer proposals. [9, 27]. At this point, it should not matter which phase sensor is used, and in fact, the same results should be obtained using any other sensor (i.e., H-S) adapted to measure with a similar resolution, perhaps in smaller areas within the pupil to be able to achieve this lateral resolution.

In this small sample of 5 healthy eyes, an optical structure in the 200-300 nm range was clearly revealed in all cases. Since the phase map represents the integration of all ocular structures, it is still too early to say exactly where this structure comes from. The next step will be the construction of a prototype capable of measuring in real time (we have had difficulties in getting a camera capable of measuring in real time with the characteristics of the one used.) In this way, in combination with cycloplegia or not or by measuring pseudophakic patients, it could be estimated whether this structure comes from the tear, the lens, the cornea, other ocular media, or the retina. However, in the repeatability measurements for the real eyes, this pattern was consistent among all measures, suggesting that it did not come from the tear. When a high-pass filter was applied, another series of details were revealed, different for each eye, for which we still do not have a clear explanation. Especially striking is the case of the most myopic eye, #MVO, where a series of parallel and vertical lines appeared that cross the pupil.

The scope of this discovery is unknown. Most likely, it will take years of clinical studies to learn how to interpret this new information, associate it with different pathologies or conditions, and determine whether it has a real clinical application or is just a curiosity. The applications of ocular aberrometry are ubiquitous: they include refractive surgery (i.e., LASIK) [28], the diagnosis and monitoring of diseases [29], myopia control [30], intraocular lenses [31], light- adjustable intraocular lenses [32], objective refraction [33], intraoperative aberrometry [34], and vision science [35]. Additionally, combining a wavefront sensor with a compensating element, such as a liquid crystal spatial light modulator, has dramatically improved in vivo observations of the human retina [36]. It has also enabled the possibility of improved visual perception, so-called super-vision, which is a visual acuity far beyond the normal values [37]. Although all of these are potential applications of this finding, it would be pure speculation to say at this point that it will have a significant impact on any of them (however, we are optimistic about it).

It is important to note that we are not using a commercial H-S sensor built expressly for ophthalmic measurements but a general-purpose sensor. Additionally, resampling the high-resolution phase map to the H-S resolution may not exactly represent what an H-S sensor would detect. We believe that the small differences between the measurements of the two sensors can be justified in this way. While different approaches could be taken to improve this aspect, especially regarding interpolation, it would be difficult to justify this, as such approaches would appear purposefully made to make the results appear accurate. For this reason, we preferred to perform a simple bilinear interpolation for the comparisons. We believe that the existence of differences between the two sensors after an exhaustive validation of WFPI is also valuable information, and it may be behind the typical errors of H-S sensors applied in ophthalmology. [38-41]. However, analysing in detail what the H-S is not capable of detecting would deviate too much from the main objective of this work, which is to show the phase map of the human eye with high lateral resolution, and it will be the subject of future studies.

## Conclusion

In this work, we showed that when the optics of the human eye are measured at a lateral resolution of 8.55 µm, a series of optical substructures with a pattern of peaks and valleys that appear in an amplitude range of 200 to 300 nm are revealed. Furthermore, the phase map of the human eye was shown to be rich in high frequencies when a high-pass filter is applied to the result at a high resolution. This is the first time this finding has been described, and it could have a significant impact on our understanding of some fundamental mechanisms of human vision as well as outstanding utility in certain fields of ophthalmology.

## Data Availability

The data used to generate figure 6 are included for anyone to use at https://bitbucket.org/wooptix/wfpi_data. The rest of the data generated or analysed during this study are included in this published article. These additional data are available from the corresponding authors upon reasonable request.

## Acknowledgements

We thank José Luis Hernández-Afonso for his administrative and logistical assistance. We thank Jan Gaudestad for a critical review of the manuscript.

## Author contributions

S.B.G. contributed to all stages of the project. J.M.T.S. contributed to all stages of the project. M.V.O. contributed to all stages of the project except the development of the WFPI algorithm. O.C.G. contributed as the main computer engineer. M.S.C. contributed as the main electronic engineer. A.R.V. contributed as the main mechanical engineer. S.C. contributed to the development of the WFPI algorithm. R.O.G. contributed to the processing software and hardware control. J.M.H. contributed to the hardware control. O.G.C. contributed to the hardware control. J.G.M.H. contributed to the processing software. D.G. contributed to the interpretation of data. J.T.H. contributed to the interpretation of data. J.M.R.R. contributed to the development of the WFPI algorithm and to the conception and design of the work.

## Additional information

The data used to generate figure 6 are included for anyone to use at https://bitbucket.org/wooptix/wfpi_data. The rest of the data generated or analysed during this study are included in this published article. These additional data are available from the corresponding author upon reasonable request.

### The authors declare the following competing interests

S.B.G., employment. J.M.T.S., employment. M.V.O., employment. O.C.G., employment. M.S.C., employment. A.R.V., employment. S.C., employment. R.O.G., employment. J.M.H., employment. O.G.C., employment. J.G.M.H., employment. D.G., personal financial interests. J.T.H., personal financial interests. J.M.R.R., personal financial interests.

